## Supplementary material for "Population risk factors for severe disease and mortality in COVID-19: A global systematic review and meta-analysis": S1_Table

**Supplementary Table 1.** Systematic literature review search terms and strategy.

**Search strategy (PICO)**

1. COVID-19 [Supplementary Concept]

2. (ventilator OR ICU OR intensive care OR MODSOR ARDS OR severity OR prognosis OR hospitalis* OR hospitaliz* OR intubation OR ventilation OR admission* OR admitted OR "critical care" OR "critical cases"OR severe) [Title/Abstract]

3.(clinical OR symptom* OR characteristic* OR comorbidit* OR “co morbidit*” OR "risk factors" OR predict*) [Title/Abstract]

4 (pediatric* OR paediatric* OR child*)[Title/Abstract]

5. ("2020/01/01"[Date - Publication] : "3000"[Date - Publication])

1 AND 2 AND 3 AND 5 NOT 4

**Search terms for PubMed**

((((COVID-19[Supplementary Concept]) AND ((ventilator[Title/Abstract] OR ICU[Title/Abstract] OR intensive care[Title/Abstract] OR mortality[Title/Abstract] OR prognosis[Title/Abstract] “MODS”[Title/Abstract] OR ARDS[Title/Abstract] OR severity[Title/Abstract] OR prognosis[Title/Abstract] OR hospitalis*[Title/Abstract] OR hospitaliz*[Title/Abstract] OR “respiratory failure”[Title/Abstract] OR intubation[Title/Abstract] OR ventilation[Title/Abstract] OR admission*[Title/Abstract] OR admitted[Title/Abstract] OR "critical care"[Title/Abstract] OR "critical cases"[Title/Abstract] OR severe)[Title/Abstract])) AND ((clinical[Title/Abstract] OR symptom*[Title/Abstract] OR characteristic*[Title/Abstract] OR comorbidit*[Title/Abstract] OR “co morbidit*”[Title/Abstract] OR risk[Title/Abstract] OR predict*)[Title/Abstract])) NOT ((pediatric*[Title/Abstract] OR paediatric*[Title/Abstract] OR child*)[Title/Abstract])) AND (("2020/01/01"[Date - Publication] : "3000"[Date - Publication])) Filters: English Sort by: Most Recent

**Search terms for Scopus**

(TITLE-ABS-KEY ( ncov* OR coronavirus OR "SARS-CoV-2" OR covid-19 OR covid ) AND TITLE-ABS-KEY ( ventilator OR icu OR intensive AND care OR mortality OR prognosis "MODS" OR ards OR severity OR prognosis OR hospitalis* OR hospitaliz* OR "respiratory failure" OR intubation OR ventilation OR admission* OR admitted OR "critical care" OR "critical cases" ) AND TITLE-ABS-KEY ( clinical OR symptom* OR characteristic* OR comorbidit* OR "co morbidit*OR risk OR predict* ) AND NOT TITLE-ABS-KEY ( pediatric* OR paediatric* OR child* ) ) AND DOCTYPE ( ar OR re ) AND PUBYEAR > 2019
