## Supplementary material for "Population risk factors for severe disease and mortality in COVID-19: A global systematic review and meta-analysis": S1_Figure

### Age >75 years old

| Source | OR (95% CI) |
|---|---|
| Zhou2020 | 22.30 [2.62; 189.80] |
| Wang2020 | 29.94 [3.59; 249.68] |
| Okoh2020 | 3.59 [1.38; 9.34] |
| Palaiodimos2020 | 3.59 [1.00; 12.89] |
| Huang2020 | 3.82 [1.79; 8.15] |
| Masetti2020 | 10.63 [4.29; 26.34] |
| Li2020 | 2.94 [1.14; 7.58] |
| Gupta2020 | 5.36 [3.20; 8.98] |
| Total (fixed effect) | 5.14 [3.75; 7.05] |
| Total (random effects) | 5.57 [3.10; 10.00] |

Heterogeneity: $\chi^2_7 = 9.71$ ($P$ = .21), $I^2 = 28\%$

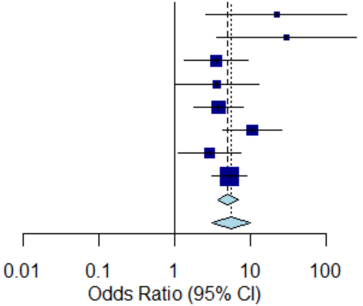


### Male

| Source | OR (95% CI) |
|---|---|
| Wang2020 | 7.22 [1.30; 40.10] |
| Palaiodimos2020 | 2.77 [1.25; 6.14] |
| Huang2020 | 2.10 [1.12; 3.94] |
| Gupta2020 | 1.50 [1.19; 1.89] |
| Total (fixed effect) | 1.66 [1.35; 2.04] |
| Total (random effects) | 2.19 [1.23; 3.89] |

Heterogeneity: $\chi^2_3 = 5.69$ ($P$ = .13), $I^2 = 47\%$

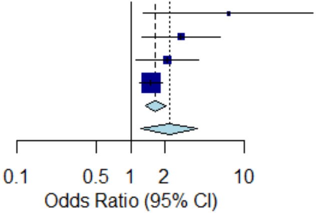
