## Supplementary material for "Population risk factors for severe disease and mortality in COVID-19: A global systematic review and meta-analysis": S2_Figure

### Diabetes

| Source | OR (95% CI) |
|---|---|
| Palaiodimos2020 | 1.16 [0.55; 2.45] |
| Gupta2020 | 1.14 [0.91; 1.43] |
| Total (fixed effect) | 1.14 [0.92; 1.42] |
| Total (random effects) | 1.14 [0.92; 1.42] |

Heterogeneity: $\chi^2_1 = 0.00$ ($P = .97$), $I^2 = 0\%$

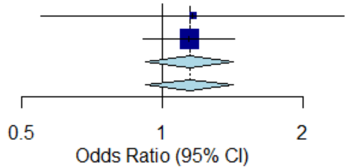


### Hypertension

| Source | OR (95% CI) |
|---|---|
| Wang2020 | 1.10 [0.26; 4.65] |
| Huang2020 | 1.26 [0.68; 2.33] |
| Gupta2020 | 1.06 [0.83; 1.35] |
| Total (fixed effect) | 1.09 [0.87; 1.36] |
| Total (random effects) | 1.09 [0.86; 1.37] |

Heterogeneity: $\chi^2_2 = 0.26$ ($P = .88$), $I^2 = 0\%$

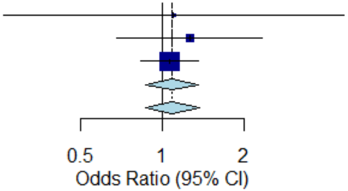


### Active cancer

| Source | OR (95% CI) |
|---|---|
| Dai2020 | 2.17 [0.81; 5.81] |
| Gupta2020 | 2.15 [1.35; 3.42] |
| Total (fixed effect) | 2.15 [1.41; 3.28] |
| Total (random effects) | 2.15 [1.41; 3.28] |

Heterogeneity: $\chi^2_1 = 0.00$ ($P = .99$), $I^2 = 0\%$

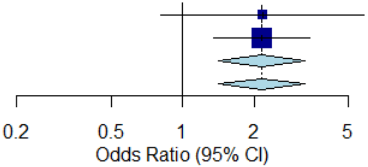
