## Supplementary figures and images for "Population risk factors for severe disease and mortality in COVID-19: A global systematic review and meta-analysis"

### S3_Figure

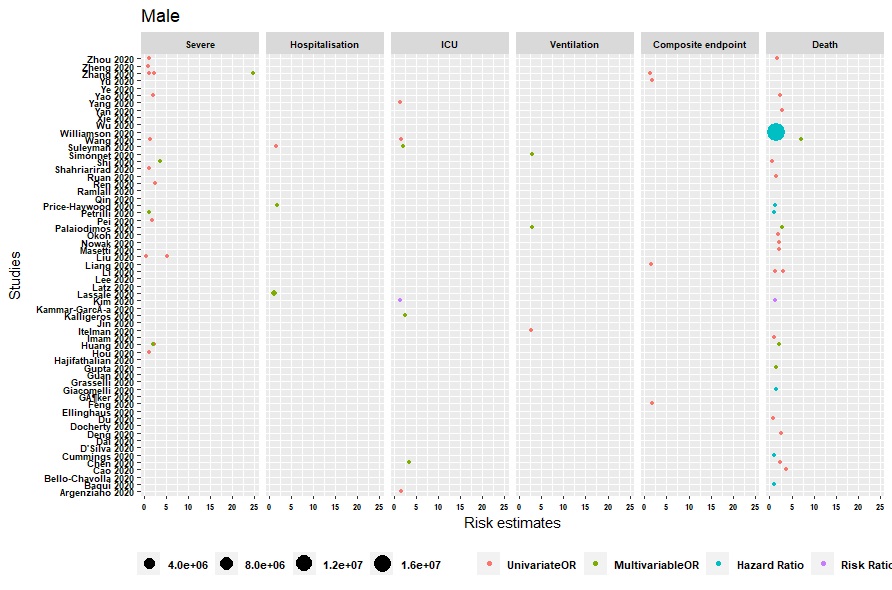

### S4_Figure

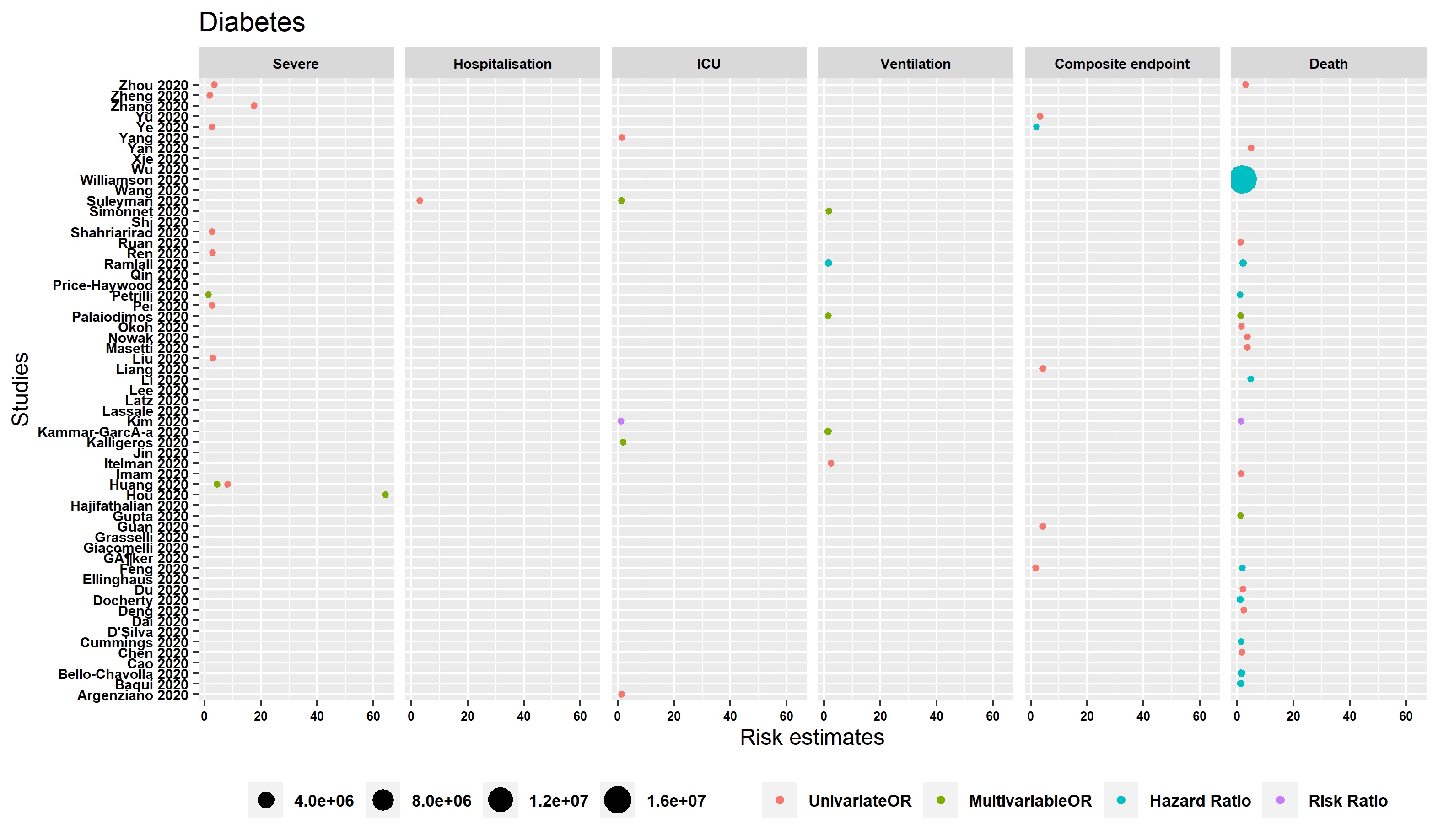

### S5_Figure

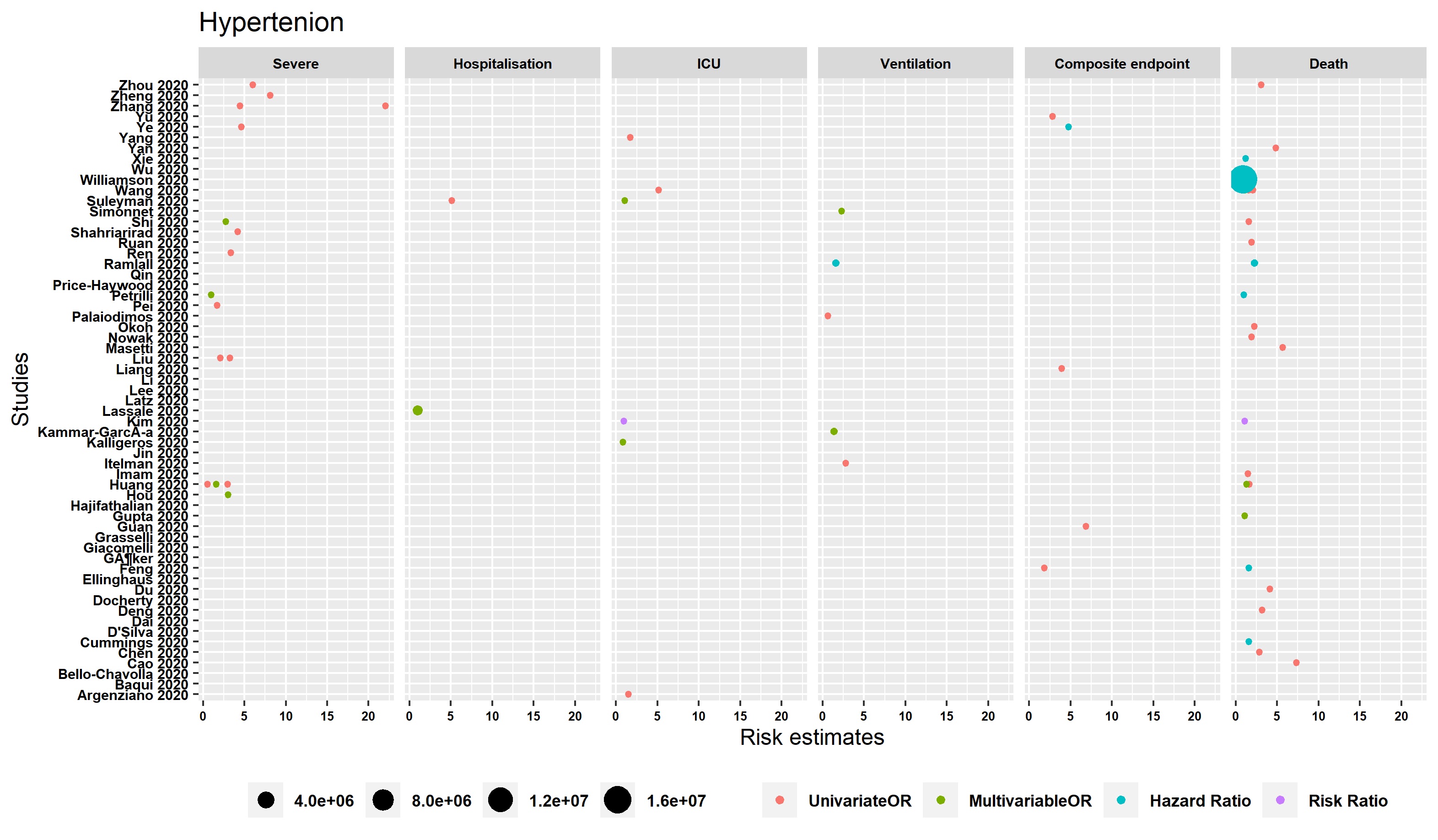
